# Risk Factors for Traumatic Brain Injury in Men and Women

**DOI:** 10.1101/2022.12.29.22284004

**Authors:** Basheer Abdullah Marzoog, Ekaterina Vanichkina

## Abstract

**Objectives:** The study covers the current status of TBI and provides a statistical recommendation to health organizations in the world.

**Design:** A retrospective analytical study. The descriptive results showed that a difference exists between males and females and for these reasons the sample was divided into two groups according to the primary descriptive statistics. The first group included 212 (31.59 %) females, while the second group included 459 (68.40%) males.

**Setting:** Assess the risk factors for TBI in both sexes and compare them with the results of international studies.

**Participants:** The study involved 671 patients for the period from 07/01/2017-17/12/2018. The primary data were collected from the republic hospital.

**Interventions:** Retrospectively analysed these patients using T test, one- and two-way ANOVA test, and the Pearson correlation test using the Statistica 12 program. The study divided into male and female by using the ROC and AUC values.

**Main Outcome Measures:** The most frequently reported cause of TBI in men and women is domestic accidents. Men are affected by TBI in early life compared to women.

**Results:** The mean age of the men is 44.41 years (Std. error 0.75). The mean age of the women is 49.50 years (Std. error 1.38). In the male group, 354 (77.12 %) patients live in the city and 105 (22.87%) live in the village. However, in the female group, 170 (80.18%) patients live in the city and 42 (19.81%) live in the village. In the men group, 172 (37.47%) patients had TBI due to domestic accidents. The most frequently reported etiology of TBI in women is domestic accident, reported in 122 (57.54 %). In female group, the mean age of patients with domestic accident associated TBI is 53.76 years (Std. error 1.85). In male group, the mean age of patients with domestic accident associated TBI is 50.74 years (Std. error 1.09). Total hospitalization days of the patients were associated with the age of the patients, r= 0.12. Where men are hospitalized longer than women, t value -2.261, p < 0.024. In the male and female groups, there is a direct correlation between age and the total hospitalization days in the male group, r=0.173; r=0.148, respectively.

**Conclusion:** The most frequently reported cause of TBI in men and women is domestic accidents. Men are affected by TBI in early life compared to women.

## Background

Traumatic brain injury is reported in multiple studies, and it is suggested that gender plays an important role in post-TBI outcomes [1]. A recent multicenter study concluded that gender plays an important role in the severity of TBI using the Glasgow coma scale (GCS) score [2]. Traumatic brain injury can be contusion and concussion, and penetrating wound injury as well as anoxic brain injuries.

Several factors play a role in the determination of the risk of TBI in men and women, such as age, hormonal status, and the presence of poor health care practice [3,4]. The classical causes of TBI include car accidents, falls from hills, sport associated injuries, and domestic violence [5]. The domestic violence is the primary cause of TBI and post TBI outcomes and complications [6].

Traumatic brain injury clinical manifestations are severity-dependent and age-dependent. Various sensory symptoms, physical symptoms, and cognitive, behavioral, or mental symptoms [7].

The dramatic increase in the number of traumatic brain injuries exposes the community to a high prevalence of disability associated with TBI associated disability[8]. Therefore, TBI is an emergency and requires urgent action to reduce the incidence rate of TBI occurrence [9]. The study sought to determine the risk factors most frequently reported for TBI occurrences.

## Methods

A retrospective analytical study involved 671 patients for the period of 07/01/2017-17/12/2018. The primary data were collected from the republic hospital for the past 2 year and retrospectively analyzed. The descriptive results showed that a difference exists between males and females and for these reasons the sample was divided into two groups according to the primary descriptive statistics. The first group included 212 (31.59 %) females, while the second group included 459 (68.40%) males. The consent of the patients has been taken for scientific purposes to analyze and publish the results of the study. The study divided into male and female by using the ROC and AUC values. (*Figure 1)*

**Figure 1.**
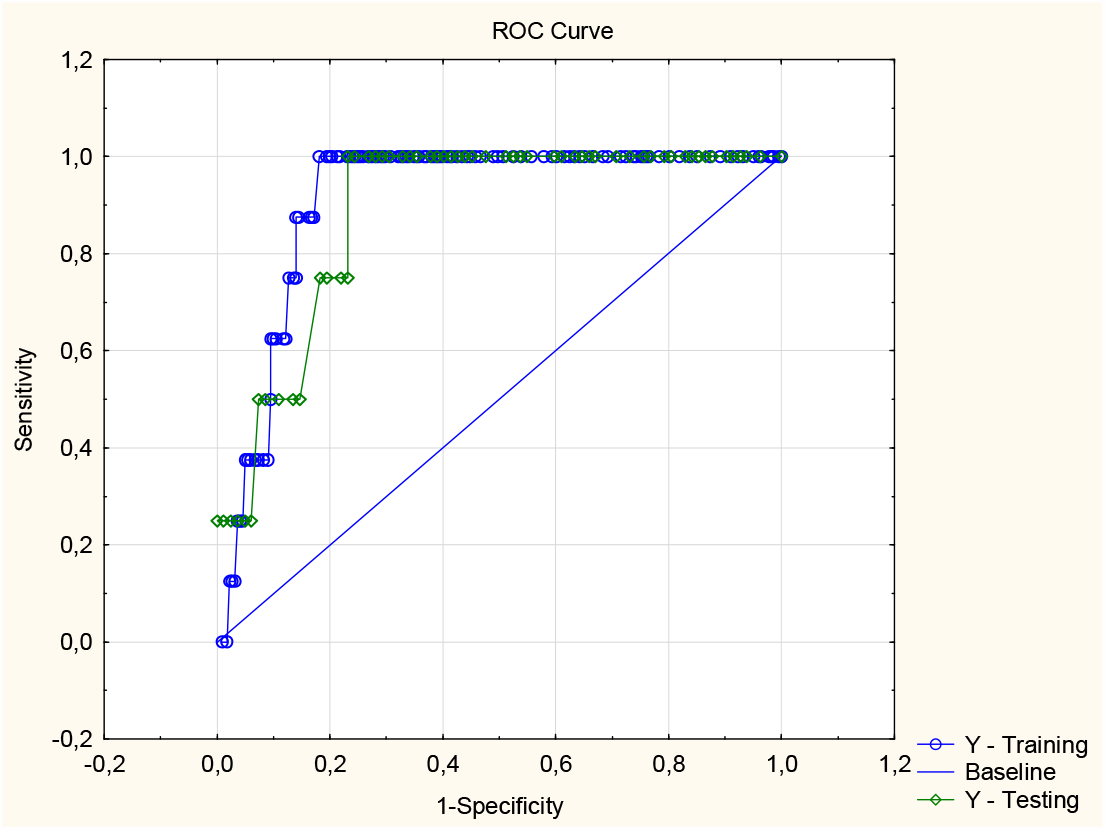
The results of ROC and AUC analysis on which the sample has been divided.

For statistical analysis, the T test, one- and two-way ANOVA test, and the Pearson correlation test were used using the Statistica program. (StatSoft, Inc. (2011). STATISTICA (data analysis software system), version 10. www.statsoft.com.).

## Results

The descriptive statistical analysis showed that the mean age of the participants is 46.016 years old (min; max: 15.00; 92.00). The mean hospitalization days ranged from 0.00 - 49.00 days (mean; Std. Error: 12.75; 0.26). The study involved 212 (31.59 %) women and 459 (68.40%) men. The demographic distribution of the sample included 524 (78.09 %) who live in the city and 147 (21.90%) who live in the village. The frequency of traumatic brain injury (TBI) due to alcohol-related brain injury is observed in 139 (20.71%) patients, due to criminal injury 75 (11.17%) patients, due to domestic accident 294 (43.81%) patients, due to work injury 16 (2.38%) patients, due to motor vehicle injury 111 (16.54 %) patients, due to postoperative haematoma 2 (0.29%) patients, due to convulsive syndrome 7 (1.043%) patients, due to sports injury 2 (0.29%) patients, due to unknown reason 25 (3.72%) patients. Number of the patients required blood transfusion is 3 (0.44%) and 668 (99.55%) required only conservative therapy.

Of 671, the need for surgical treatment was observed in 198 (39.91%) patients, missing data in 181 (26.97%), and 292 (43,51%) did not require surgical intervention. Out of 198, 125 (63.13%) patients required surgical debridement of wounds. 42 (21.32%) patients required scull trepanation with removal of acute subdural hematoma. 6 (3.03 %) patients required scull trepanation and removal of acute epidural haematoma. 8 (4.12%) patients required scull trepanation and removal of chronic subdural hematoma. 4 (2.03 %) patients required scull trepanation and removal of acute intracerebral haematoma. 3 (1.54 %) patients required scull trepanation and osteoplasty of fracture. 3 (1.52 %) patients required osteoplasty of fracture. 4 (2.03%) patients required Bulau pleural drainage. 1 (0.50 %) patient required surgical debridement of wounds and pleural drainage by Bulau. 2 (1.01%) patients required scull trepanation and removal of acute subdural haematoma with removal of acute intracerebral hematoma.

The mean age of patients affected by TBI who live in the city is 47.101 years. The mean age of the patients affected by TBI who live in the village is 42.14 years. The people who live in the village are at increased risk of TBI, t value 3.02971, p< 0.002.

Total hospitalization days of the patients were associated with the age of the patients, r= 0.12. Where men are hospitalized longer than women, t value -2.26, p < 0.024.

Mean age of patients with alcohol-related brain injury is 46.079 years (Std. error 1.17). Mean age of patients with domestic accident is 50.74 years (Std. error 1.098). Mean age of patients with motor vehicle-related injury is 38.072 years (Std. error 1.68). The mean age of patients with criminal injury is 37.94 years (Std. error 1.59). The mean age of patients with work injury is 41.75 years (Std. error 3.081). The mean age of patients with convulsive syndrome is 50.142 years (Std. error 5.65). The mean age of patients with postoperative haematoma is 55.00 years (Std. error 2.00). Mean age of patients with sports injury is 26 years (Std. error 0.00). The mean age for patients with TBI of unknown reasons is 52.04 years (Std. error 2.77).

In terms of gender-dependent risk factors, men are frequently exposed to domestic accidents as a risk factor for the development of TBI. In the men group, 172 (37.47 %) patients had TBI due to domestic accidents. However, the mean hospitalization days of these patients was 13.30 (Std. error 0.56).

The mean age of the men is 44.41 years (Std. error 0.75). The mean age of the women is 49.50 years (Std. error 1.38). Indicates that men are affected by TBI at an early age compared to women, t value 3.50, p <0.0004. (Figure 2)

**Figure 2.**
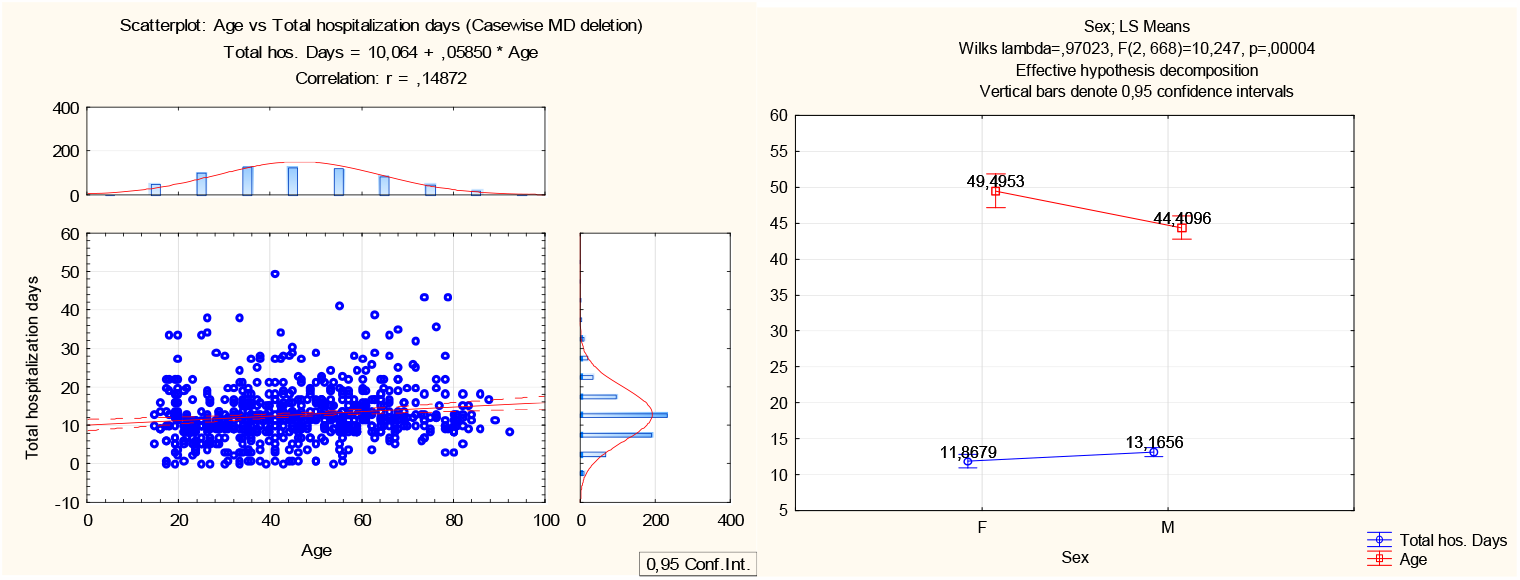
Graphical presentation of the direct association between the total hospitalization days and the age and the statistically significant difference in total hospitalization days between men and women sex of the patients.

In the male group, 354 (77.12 %) patients live in the city and 105 (22.87%) live in the village. However, in the female group, 170 (80.18%) patients live in the city and 42 (19.81%) live in the village.

The most frequently reported etiology of TBI in women is domestic accident, reported in 122 (57.54 %). However, the most frequently reported cause of TBI in males is domestic accident too. (Figure 3)

**Figure 3.**
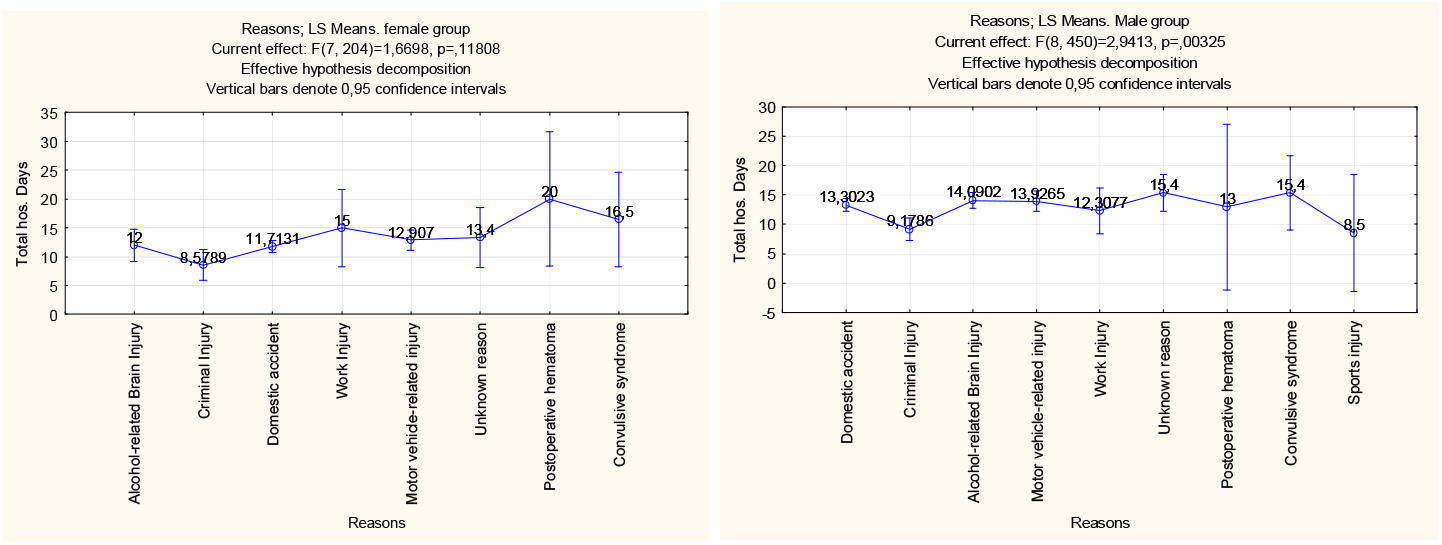
Total hospitalization days and etiology in men and women, respectively (in women statically, no significant different).

In the male group, the frequency of domestic accident-associated TBI seen in 172(37.47 %) patients, alcohol-related brain injury-associated TBI in 122 (26.57 %) patients, motor vehicle-related injury associated TBI in 68 (14.81 %) patients, criminal injury associated TBI in 56 (12.20 %) patients, work injury associated TBI in 13 (2.83 %) patients, convulsive syndrome-associated TBI in 5 (1.089 %) patients, sports injury associated TBI in 2 (0.43%) patients, postoperative haematoma-associated TBI in 1 (0.21 %), and unknown reason in 20 (4.35 %) patients. In the female group, domestic accidents were reported in 122 (57.54 %) patients, motor vehicle injury reported in 43 (20.28%) patients, criminal injury reported in 19 (8.96%) patients, alcohol-related brain injury reported in 17 (8.018%) patients, work injury reported in 3 (1.41 %) patients, convulsive syndrome reported in 2 (0.94340 %) patients, postoperative hematoma reported in 1 (0.47%) patient and unknown reason seen in 5 (2.35 %) patients.

In the male group, 458 (99.78 %) patients required conservative therapy and 1 (0.21%) required blood transfusion. However, in the female group, 210 (99.05 %) required conservative surgery and 2 (0.94 %) were treated without conservative therapy.

In male group, the mean age of patients with alcohol-related brain injury is 46.079 years (Std. error 1.17). The mean age of patients with TBI associated with domestic accidents is 50.74 years (Std. error 1.098). The mean age of patients with motor vehicle injury-associated TBI is 38.072 years (Std. error 1.68). Mean age of patients with criminal injury associated TBI is 37.94 years (Std. error 1.59). Mean age of patients with work injury associated TBI is 41.75 years (Std. error 3.081). The mean age of patients with convulsive syndrome associated TBI is 50.14 years (Std. error 5.65). The mean age of patients with sports injury-associated TBI is 26 years (Std. error 0.00). The mean age of patients with postoperative haematoma-associated TBI is 57 years (Std. error 0.00). The mean age of patients with unknown reason associated TBI is 52.04 years (Std. error 2.77). In the female group, the mean age of patients with alcohol-related brain injury is 43.52 years (Std. error 3.63). The mean age of patients with domestic accident-associated TBI is 53.76 years (Std. error 1.85). Mean age of patients with motor vehicle-related injury associated TBI is 40.30 years (Std. error 3.069). Mean age of patients with criminal injury associated TBI is 37.94 years (Std. error 1.59). Mean age of patients with work injury associated TBI is 45.33 years (Std. error 4.17). The mean age of patients with convulsive syndrome associated TBI is 57.5 years (Std. error 5.50). The mean age of patients with sports injury-associated TBI is 26 years (Std. error 0.00). The mean age of patients with postoperative haematoma-associated TBI is 53 years (Std. error 0.00). The mean age of patients with unknown reason is 61.20 years (Std. error 6.39).

In male group, out of 459 patients with TBI, 158 (46.19%) required surgical treatment and 184 (53.80%) resolved without surgical treatment, missing data in 117 (25.54%) patients. Of 158 patients, 3 (1.89%) required fracture osteoplasty. 94 (59.49%) patients required surgical debridement of the wounds. 11 (6.96%) required scull trepanation and removal of chronic subdural haematoma. 1 (0.63%) required scull trepanation with removal of acute subdural hematoma. 38 (24.05%) required scull trepanation and removal of acute subdural hematoma. 2 (1.26%) required scull trepanation with fracture osteoplasty. 1 (0.63%) required surgical debridement of wounds with pleural drainage by Bulau. 5 (3.16%) required scull trepanation with removal of acute epidural haematoma. 3 (1.89%) required scull trepanation with removal of acute intracerebral haematoma. 2 (1.26%) required scull trepanation and removal of acute subdural haematoma, as well as removal of acute intracerebral hematoma. In the female group, out of 212 patients, 40 (18.87%) patients required surgical treatment, missing data in 64 (30.18%) patients. Of 40, 31 (77.50%) patients required surgical debridement of wounds, 4 (10.00%) required scull trepanation with removal of acute subdural haematoma, 1 (2.50%) required scull trepanation with removal of acute epidural haematoma, 2 (5.00%) required scull trepanation with removal of chronic subdural haematoma, 1 (2.50%) required scull trepanation with removal of acute intracerebral haematoma, and 1 (2.50%) required scull trepanation with fracture osteoplasty. (*Table 1*)

**Table 1:** Non parametric characteristics of the sample.

| Factor | Index | Group M ( ) | Group F ( ) | X<sup>2</sup> | P value |
| --- | --- | --- | --- | --- | --- |
| Sex | M | 459 (100.00 %) | 0 (0.00 %) | 671.000 | <0,001 |
|  | F | 0 (0.00 %) | 212 (100.00%) |  |  |
| Living place | City | 354 (77,12 %) | 170 (80,18 %) | 0.796 | 0.373 |
|  | Village | 105 (22,87 %) | 42 (19,81 %) |  |  |
| Conservative therapy | Yes | 458 (99,78 %) | 210 ( ) | 0.373 | 0.191 |
|  |  |  | 99,05 %) |  |  |
|  | Blood transfusion | 1 (0,21 %) | 2 (0,94 %) |  |  |

In the male group, the mean hospitalization days for patients with TBI who live in the city are 12.83 days and 14.29 days for patients who live in the village. In the female group, the mean hospitalization day is 11.87 days (Std. error 0.40). However, in the female group, the mean days for TBI patients living in the city are 11.79 days and 12.19 days for patients living in the village. The Mean total hospitalization days of patients with alcohol-related brain injury is 12.00 days (Std. error 1.85). The mean total hospitalization of patients with domestic accidents is 12.64 days (Std. error 0.40). The mean total hospitalization of patients with motor vehicle-related injury is 13.53 days (Std. error 0.66). The mean total hospitalization of patients with criminal injury is 9.026 days (Std. error 0.61). The mean total hospitalization of patients with work injury is 12.81 days (Std. error 2.029). The mean total hospitalization of patients with convulsive syndrome is 15.71 days (Std. error 3.037). The mean total hospitalization of patients with sports injuries is 8.5 days (Std. error 3.50). The mean total hospitalization of patients with postoperative hematoma is 16.50 days (Std. error 3.50). The mean total hospitalization of patients for unknown reason is 15.00 days (Std. error 1.34).

In the male group, the mean total hospitalization days of patients with alcohol-related brain injury is 14.09 days (Std. error 0.62). The mean total hospitalization of patients with domestic accidents is 13.30 days (Std. error 0.56). The mean total hospitalization of patients with motor vehicle-related injury is 13.92 days (Std. error 0.94). The mean total hospitalization of patients with criminal injury is 9.17 days (Std. error 0.76). The mean total hospitalization of patients with work injury is 12.30 days (Std. error 2.47). The mean total hospitalization of patients with convulsive syndrome is 15.4 days (Std. error 4.38). The mean total hospitalization for sports injuries is 8.5 days (standard error 3.5). The mean total hospitalization of patients with postoperative haematoma is 8.5 days (Std. error 3.50). The mean total hospitalization of patients for unknown reason is 15.4 days (Std. error 1.62). In the female group, the mean total hospitalization days of patients with alcohol-related brain injury is 12.00 days (Std. error 1.85). The mean total hospitalization of patients with domestic accident is 11.71 days (Std. error 0.55). The mean total hospitalization of patients with motor vehicle-related injury is 12.90 days (Std. error 0.85). The mean total hospitalization of patients with criminal injury is 8.57 days (Std. error 0.85). The mean total hospitalization of patients with work injury is 15.00 days (Std. error 1.73). The mean total hospitalization of patients with convulsive syndrome is 15.4 days (Std. error 4.38). The mean total hospitalization days of patients with sports injury is 8.5 days (Std. error 3.5). The mean total hospitalization of patients with postoperative haematoma is 8.5 days (Std. error 3.50). The mean total hospitalization of patients for unknown reason is 13.40 days (Std. error 1.80).

In the male group, the mean age of patients with TBI who live in the city is 44.85 years and 42.92 years for patients who live in the village. Whereas, in the female group, the mean age of patients with TBI who lives in the city is 51.79 years old and 40.21 years old for patients living in the village, statically significant difference in the age of the TBI female patients, t value 3.42, p< 0.0007. (*Figure 4*)

**Figure 4.**
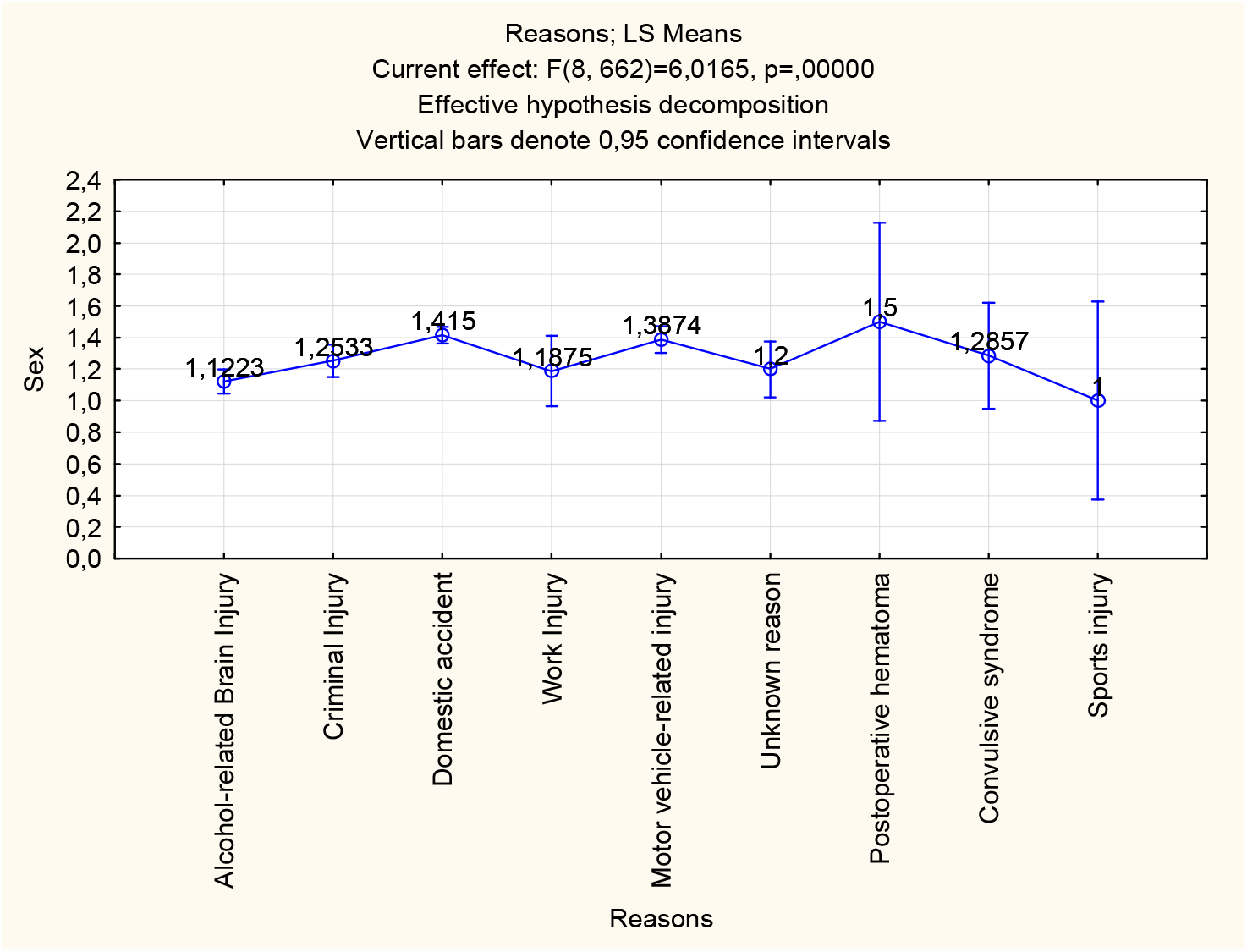
Graphical presentation of the risk markers in males and females in the republic. (The number 1 indicates males and number 2 indicates females; the nearest values to 1 demonstrates higher incidence in males and oppositely, the value nearest to 2, indicates higher incidence in female).

In the male and female groups, there is a direct correlation between age and the total hospitalization days, r=0.17; r=0.14, respectively.

## Discussion

The city population affected by TBI in older age in compare to village citizens. The total hospitalization days are longer in men than in women. Longer hospitalization days are observed in older people. Living in the village posses’ higher risk of TBI, probably due to the lack of primary health care facilities. Men are affected by TBI in early life compared to women. The frequency of TBI causes is associated with the age of the patients. Consuming alcohol is a huge risk factor for TBI. Poor hygienic practice and bad habits such as heavy alcohol drinking are avoidable risk factors.

The limitation of a study is the absence of the control group to compare the incidence rate between men and women. The outcomes of post TBI depend on the gender [2,10–18]. Different studies suggested that age plays a central role in increasing the risk of TBI [19]. In particular, women under 45 years of age and older than 65 years of age are at increased risk than men of the same age [10]. Furthermore, being a male, the risk factor for TBI is two times higher as being a woman. However, a noticeable increase in the incidence rate of TBI in women is observed compared to previous studies [20,21]. Probably associated with women’s increase in the participation in different kinds of contact sports (such as rugby), military services, and home violence [5,22–25].

## Conclusions

The most frequently reported cause of TBI in men and women is domestic accidents. Men are affected by TBI in early life compared to women. The people who live in the village are at increased risk of TBI. Gender plays a crucial role in the severity and incidence rate of post-TBI outcomes and complications. The results of our study expand the knowledge in the field of traumatic brain injury risk factors in a gender-dependent manner. Also, the results of our study are in constant with the results of the previous studies [15,17,26,27].

## Data Availability

All data produced in the present study are available upon reasonable request to the authors

## List of abbreviations

TBI: traumatic brain injury

## Declarations

### 1. Ethics approval and consent to participate

The study approved by the National Research Mordovia State University, Russia, from “Ethics Committee Requirement N8/2 from 30.06.2022”.

### 2. Consent for publication

Written informed consent was obtained from the participants for publication of study results and any accompanying images.

### 3. Availability of data and materials

applicable on reasonable request.

### 6. Authors’ contributions

MB is the writer, researcher, collected and analyzed data, and revised the manuscript, EV collected the primary data from the hospital. All authors have read and approved the manuscript.

## 7. Acknowledgments

not applicable

## 8. Authors’ information

**Basheer Abdullah Marzoog**, Research Center «Digital Biodesign and Personalized Healthcare», I.M. Sechenov First Moscow State Medical University (Sechenov University), 119991 Moscow, Russia; postal address: Russia, Moscow, 8-2 Trubetskaya street, 119991. (, +79969602820). ORCID: 0000-0001-5507-2413. Scopus ID: 57486338800. **Ekaterina Vanichkina**, National Research Ogarev Mordovia State University. ORCID: 0009-0006-3015-2306

9. The paper has not been submitted elsewhere

